# The effect of sedentary behaviour and physical activity on 1719 diseases: a Mendelian randomisation phenome-wide association study (MR-PheWAS)

**DOI:** 10.64898/2026.04.10.26350507

**Authors:** Jiayao Xu, Richard M.A. Parker, Kirsty Bowman, Gemma L. Clayton, Deborah A. Lawlor

**Author notes:** Corresponding author: Jiayao Xu, Population Health Sciences, Bristol Medical School, University of Bristol, Bristol, BS1 5DS, United Kingdom. Joint senior authors (Gemma L. Clayton, Deborah A. Lawlor).

## Abstract

**Background:** Higher levels of sedentary behaviour, such as leisure screen time (LST), and lower levels of physical activity are associated with diseases across multiple body systems which contribute to a large global health burden. Whether these associations are causal is unclear. The primary aim of this study is to investigate the causal effects of higher LST (given greater power) and, secondarily, lower moderate-to-vigorous intensity physical activity (MVPA), on a wide range of diseases in a hypothesis-free approach.

**Methods:** A two-sample Mendelian randomisation phenome-wide association study was conducted for the main analyses. Genetic single nucleotide polymorphisms (SNPs) were first selected as exposure genetic instruments for LST (hours of television watched per day; 117 SNPs) and MVPA (higher vs. lower; 18 SNPs) based on the genome-wide significant threshold (p < 5×10^-8^) from the largest relevant genome-wide association study (GWAS). For disease outcomes, we used summary results from FinnGen GWAS, including 1,719 diseases defined by hospital discharge International Classification of Diseases (ICD) codes in 453,733 European participants. For the main analyses, we used the inverse-variance weighting method with a Bonferroni corrected p-value of p ≤ 3.47×10^-4^. Sensitivity analyses included Steiger filtering, MR-Egger and weighted median analyses, and data from UK Biobank were used to explore replication.

**Findings:** Genetically predicted higher LST was associated with increased risk of 87 (5.1% of the 1,719) diseases. Most of these diseases were in musculoskeletal and connective tissue (n=37), genitourinary (n=12) and respiratory (n=8) systems. Genetic liability to lower MVPA was associated with six diseases: three in musculoskeletal and connective tissue and genitourinary systems (with greater risk of these diseases also identified with higher LST), and three in respiratory and genitourinary systems. Sensitivity analyses largely supported the main analyses. Results replicated in UK Biobank, where data available.

**Conclusions:** Higher levels of sedentary behaviour, and lower levels of physical activity, causally increase the risk of diseases across multiple body systems, making them promising targets for reducing multimorbidity.

## Introduction

Higher levels of sedentary behaviour, such as leisure screen time (LST), and lower levels of physical activity have been associated with increased risks of many diseases, affecting most systems, including cardiovascular, respiratory, musculoskeletal, reproductive, neurological, as well as mental health diseases and several cancers [1–6]. Based on the Global Burden of Disease Study, a substantial number of deaths and disability-adjusted life-years (DALYs) due to chronic diseases (e.g., cardiovascular diseases, type 2 diabetes) may be attributable to lower levels of physical activity [7, 8]. A meta-analysis of prospective cohort studies showed that higher levels of physical activity prevent death [9].

Most of the evidence on the relationship between physical activity, or sedentary behaviour, and diseases is from conventional multivariable regression analyses. Whilst this work suggests that high levels of sedentary behaviour and low levels of physical activity might be an important determinant of multimorbidity, the results from such association studies can be influenced by residual confounding or reverse causation, rather than reflecting causal effects of the exposures on diseases. With multimorbidity increasing globally – in a manner not fully explained by the ageing population – there is growing awareness of a need to improve on our currently poor understanding of it to better prevent and treat it [10]. If lower levels of physical activity and higher levels of sedentary behaviour do cause many diseases across multiple systems, greater environmental and individual support to increase physical activity and reduce sedentary behaviour could reduce multimorbidity and improve overall population health outcomes.

Mendelian randomization (MR), which typically uses genetic variants as instrumental variables for exposures, is widely used to explore unconfounded effects of exposures on outcomes in observational data, under specific assumptions [11]. As genetic variants are randomly assorted at conception, they are less likely to be affected by confounding and reverse causation than conventional multivariable regression [12]. MR phenome-wide association studies (MR-PheWAS) are an extension of MR that test the effects of one or more exposures on a wide range of diseases, selected in a hypothesis-free way [13]. This approach has been used to explore potential causes of multimorbidity such as insomnia [14] and alcohol consumption [15].

In a search for previous MR studies investigating the effects of physical activity or sedentary behaviour on diseases, we identified 155 studies up to March 2025 (Supplementary Table S1); of these, 95 focused on physical activity, 24 on sedentary behaviour and 36 on both. Each of these studies explored effects on a limited number of disease types, specifically those of the musculoskeletal and connective tissue system, cardiovascular diseases, cancers, nervous system, endocrine, nutritional and metabolic diseases and mental and behavioural disorders. We found a published MR-PheWAS that investigated the effects of physical activity on a number of gastrointestinal diseases only, but it did not examine diseases of other systems. Thus, whilst MR studies to date provide some evidence to support causal effects of lower sedentary behaviour or higher physical activity on a limited number of widely studied diseases, none have undertaken a hypothesis-free approach [16] across diseases occurring in all systems to identify novel effects and determine the extent to which these might cause multimorbidity.

The primary aim of this study was to explore the effects of higher sedentary behaviour and, secondarily, lower physical activity on diseases across multiple systems.

## Methods

Figure 1 summarises the steps in our MR PheWAS. First, we identified independent genetic variants to instrument LST and moderate-to-vigorous intensity physical activity (MVPA). We then explored the associations of those single nucleotide polymorphisms (SNPs) with disease outcomes in the discovery data (FinnGen [17]), ensuring that the alleles were harmonised to match those used as the genetic instruments. For main MR analyses, we used inverse variance weighted (IVW) method, with 1,719 diseases included within broad ICD-10 chapters. Whilst these diseases are distinct, they are not all independent within chapters, so a Bonferroni corrected p-value of p ≤ 0.05/144 (3.47×10^-4^) was used based on the total number of ICD-10 blocks (n=144), the tier of disease classification below that of chapter. Effects for LST and MVPA that passed the threshold were taken forward for MR sensitivity analyses to test bias, and also for replication in UK Biobank [18], for which we repeated the main IVW analyses for diseases with available GWAS data. Results were defined as replicating if the direction of effect in UK Biobank was the same as that in the FinnGen discovery analyses, and p ≤0.05.

**Figure 1.**
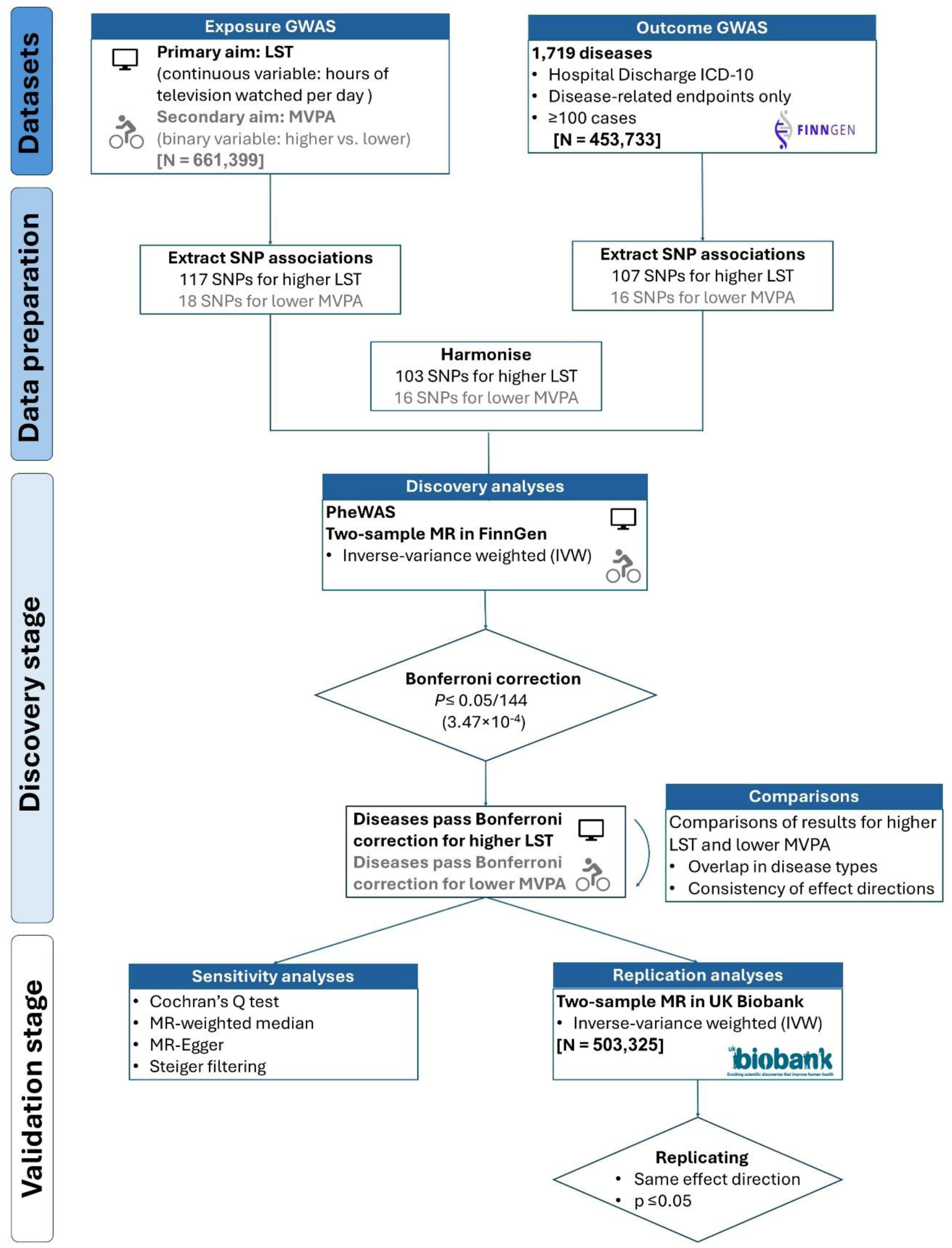
Summary of study design: Notes: LST: leisure screen time; MVPA: moderate-to-vigorous intensity physical activity during leisure time, where higher *vs.* lower MVPA equates to engaging in moderate-to-vigorous intensity physical activity for 20 minutes or more per week or two times or more per week (higher, coded as 0) *vs.* engaging in moderate-to-vigorous intensity physical activity for less than 20 minutes per week or once or less per week (lower, coded as 1).

Given people with high levels of LST are likely to have low levels of MVPA [19], *a priori* diseases affected by higher levels of LST are likely to be similarly affected by lower MVPA. Because we have more statistical power to detect effects of LST, a continuous measure (see below), than for lower MVPA, which was available only as a binary variable in the GWAS [20], we considered the estimation of LST effects to be the primary aim, and compared the extent to which the MVPA results overlapped with those for LST. For those MVPA estimates that did not overlap, we explored whether the direction of the effects was consistent, but imprecise (e.g. failing to meet the p-value threshold due to lower statistical power) by testing directional concordance.

### Exposure genetic instrument genome-wide association study (GWAS) and genetic variant selection for physical activity and sedentary behaviours

We used SNPs identified from the largest and most recent GWAS available for LST and moderate-to-vigorous intensity physical activity (MVPA). This GWAS meta-analysed results from 51 studies comprising 661,399 European adults, with UK Biobank contributing 68% of the sample [21] (Supplementary Table S2). Associations were adjusted for age, age^2^ and the top 10 ancestral principal components (PCs) (Supplementary Material S1).

In this GWAS, LST was analysed as a continuous measure of hours of television watched per day (24 hours). MVPA was analysed as a dichotomous exposure, with “higher MVPA” defined as ≥20 minutes versus <20 minutes per week for studies reporting duration (n=35), and ≥2 versus <2 times per week for studies reporting frequency (n=3) [13]. In all studies contributing to the GWAS, both exposures were assessed by self-report, with the specific questionnaires listed in Supplementary Table S2.

To be consistent with LST, we coded “higher MVPA” as 0 (reference category) and “lower MVPA” as 1. We initially selected 7,462 SNPs for LST and 1,374 SNPs for MVPA based on genome-wide significance threshold (p < 5×10^-8^). To ensure that only independent SNPs were included as genetic instrumental variables, we identified correlated SNPs defined as linkage disequilibrium (LD) r^2^ >0.001, within a 10,000 kb window using the 1000 Genomes European reference panel [22]. Where we found groups of correlated SNPs, we selected the one with the lowest p-value. Following this, we retained 117 independent SNPs for the LST genetic instrument and 18 independent SNPs for the lower MVPA genetic instrument (Supplementary Table S3).

### Outcome discovery and replication data

FinnGen is an open study pooling GWAS data from Finnish genetic biobanks [23] and national electronic health record data from hospital discharge registries (available since 1969) – which provide diagnoses for all inpatient and outpatient visits – and other national health registries (Finnish cancer register, cause of death register, drug purchases, drug reimbursement, and primary care register). It includes a nationally representative adult Finnish population with a median age of 63 years [24]. We used the most recent data release (R11) of disease GWAS summary results available when starting this study (August 2024). This included 453,733 participants of European ancestry. The GWAS was adjusted for sex, age, 10 ancestry PCs, FinnGen chip version and legacy genotyping batch.

From FinnGen’s list of clinical endpoints, we selected diseases identifiable in the hospital discharge registry that could be mapped to the International Classification of Diseases, Tenth Revision (ICD-10) codes, which is used in UK Biobank and therefore offered the potential to replicate findings in an independent large study. In FinnGen (discovery study), disease ICD codes were initially identified in the hospital discharge registry, and other registries were used only when an eligible disease was also recorded. We excluded 90 pregnancy-related diseases, such as gestational diabetes or pre-eclampsia, as our exposure genetic instruments are not specific to pregnancy. Diseases with fewer than 100 cases were excluded to minimize the risk of false negatives (type-2 errors) [25]. In total, we included 1,719 diseases with sample sizes ranging from 54,913 to 453,733, and numbers of cases from 100 to 165,937 (Supplementary Table S4). This included 1,590 diseases that occur in both sexes, 105 diseases that were female-specific, and 24 male-specific (Figure 2).

**Figure 2.**
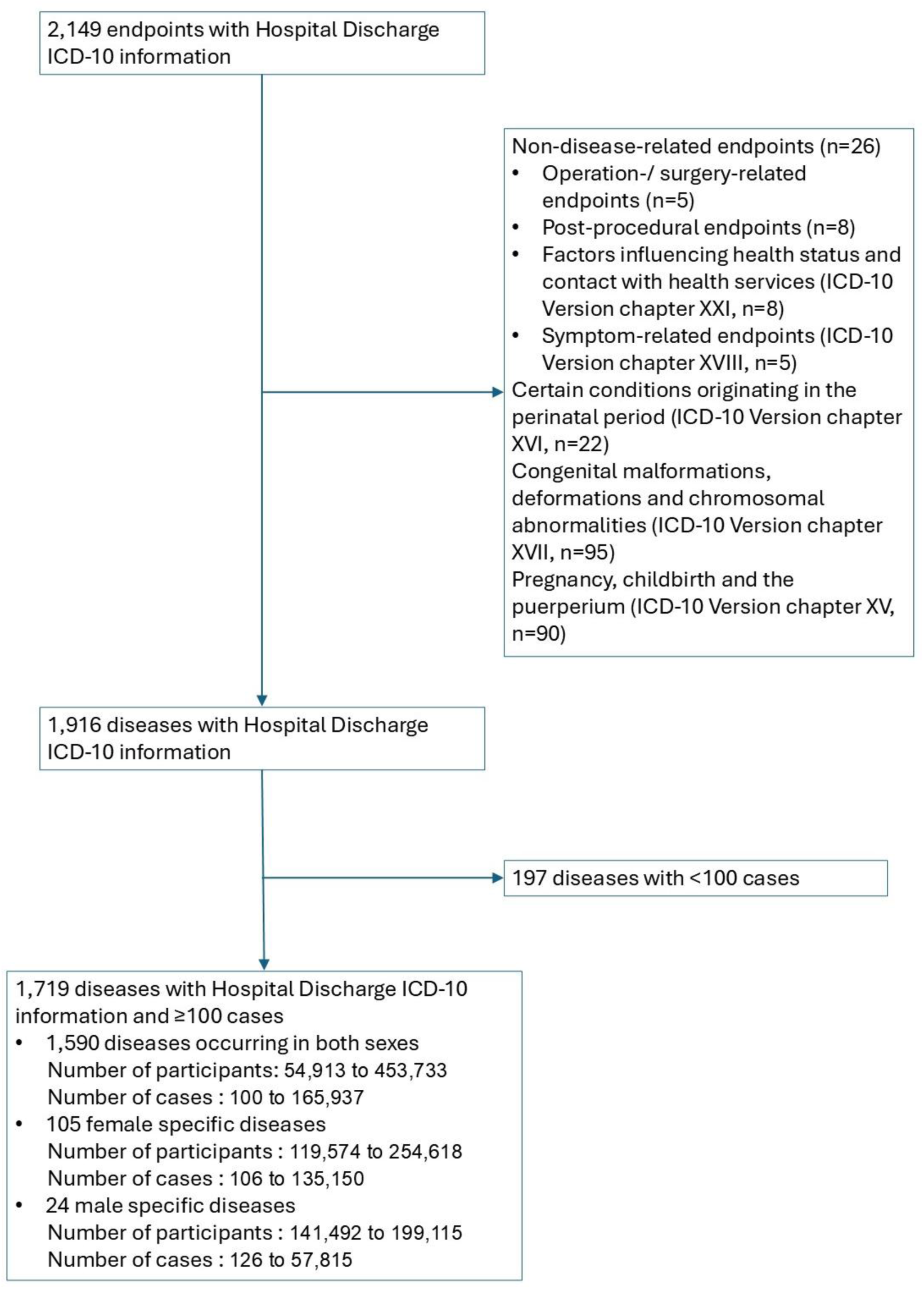
Flowchart detailing the exclusion and inclusion of diseases.

UK Biobank is a UK prospective cohort study that enrolled 503,325 adults (5.5% of those invited) aged 40 to 69, with 91% identifying as white European [18]. Participants were recruited across 22 assessment centres between 2006 and 2010. It includes genome-wide data and linked hospital episode statistics (HES), covering inpatient admissions only (no outpatient consultations), with ICD-coded diseases recorded since 1997 and continuously updated [26]. To explore replication of our FinnGen results, we used GWAS data for HES ICD-10 coded diseases in the UK Biobank, available through the Neale lab (https://www.nealelab.is/uk-biobank) [27], OpenGWAS platform (https://gwas.mrcieu.ac.uk/datasets) [28] or GeneATLAS (http://geneatlas.roslin.ed.ac.uk/) [29]. The latest UK Biobank GWAS, using disease definitions similar to FinnGen, was used for the replication analyses, based on data available up to 2018 when the GWAS analyses were conducted. The covariates adjusted for in these GWASs are listed in Supplementary Table S5 [27, 29, 30].

### Harmonisation of exposure and outcome genetic data

We searched for all of the SNPs that were included in either of the exposure genetic instrumental variables and then extracted SNP data (i.e. difference in mean LST per allele and the relevant standard error [SE] for these associations and odds ratios for lower MVPA and related SE) from FinnGen across the whole genome-wide data irrespective of p-values. Where an exposure SNP was not present in the outcome GWAS, we explored whether a proxy SNP — defined as another SNP that is highly correlated with the target SNP (LD r^2^>0.8) — could be identified. We then harmonised the outcome SNPs to ensure consistency with the allele order of the corresponding exposure SNPs and checked that the harmonisation was successful. For LST, 14 SNPs could not be harmonised because no suitable proxy was available for 10 missing SNPs and 4 SNPs were palindromic, resulting in 103 independent SNPs included in the genetic instrument. For MVPA, it was not possible to harmonise two SNPs because no suitable proxy was available for the missing SNPs; this resulted in 16 independent SNPs being included in the genetic instrument.

The same extraction and harmonisation methods were applied in the replication analyses, which were limited to SNPs for specific diseases with evidence of effects.

### Discovery Analysis

The TwoSampleMR (MR-base) R package v 0.6.4 was used to extract and harmonise outcome SNPs and to run the analyses [31]. We used the inverse-variance weighted (IVW) method for the main MR analyses. The IVW method estimates the effect of interest using an inverse variance weighted regression of SNP-outcome associations on SNP-exposure associations, with the regression intercept constrained to zero [32]. This restriction to a zero intercept implies that there is no bias due to unbalanced horizontal pleiotropy. Results for LST are presented as odds ratio (of the disease) per one-hour more LST per day. As the GWAS for MVPA used a binary measure, we are instead estimating the effect of genetic liability to lower MVPA, rather than the exact exposure [33]; specifically, we estimate the odds ratio (of the disease) per unit increase in the genetically predicted log-odds of lower MVPA, with higher MVPA as the reference group (i.e., ≥20 minutes of MVPA per week compared to <20 minutes per week, or ≥2 times per week compared to <2 times per week) [21]. In addition to Bonferroni corrected p-values, we also provide adjusted p-values using the Benjamini-Hochberg procedure to control the false discovery rate (FDR) at 5% [34].

### Exploring possible violations of MR assumptions and sensitivity analyses

MR has three core assumptions: 1) the genetic instruments are statistically strongly associated with the exposure in the relevant population (relevance assumption); 2) there is no confounding between the genetic instruments and disease (independence assumption); 3) the genetic instruments influence the disease(s) only through the exposure and not through any other pathway (exclusion restriction assumption) [35].

To assess these assumptions, we explored between-SNP heterogeneity tests for each exposure-outcome IVW analysis, using Cochran’s Q statistic. Between-SNP heterogeneity can occur when any of the assumptions are violated. We used the mean F-statistic across all SNPs to test the strength of the genetic instrument for LST, and the pseudo mean F statistic for MVPA [36].

Whilst genetic variants are unlikely to be influenced by conventional confounding, such as socioeconomic status and behavioural traits [12], confounding of genetic instrument-outcome associations can occur when there is population stratification, such as different ancestral groups, with different allele frequencies and risk of outcomes. To mitigate against this, we included European participants only, and all the GWAS we used were adjusted for genetic ancestral PCs.

We explored bias due to unbalanced horizontal pleiotropy by running sensitivity analyses using two pleiotropic robust methods: weighted median and MR-Egger [35]. The weighted median method can provide reliable estimates, provided that at least 50% of the weight of the instrumental variable is valid [37]. The MR-Egger method is identical to the IVW method with the exception that it does not force the intercept to be zero, meaning that the regression slope, i.e. effect estimate, is corrected for unbalanced horizontal pleiotropy and the p-value for the intercept is a test of the null hypothesis that there is no unbalanced horizontal pleiotropy.

In an additional sensitivity analysis, we explored whether any of the diseases had potential effects on LST or MVPA, generating spurious estimates of their effect on diseases, using Steiger filtering. This compares the amount of variation explained in the exposure and outcome for each SNP used in the genetic instrument. We re-ran the IVW analyses, excluding any SNPs that explained more variation in the outcome than in the exposure, to see if these were introducing bias [38].

## Results

We included 1,719 diseases in this MR-PheWAS. The mean F-statistics (pseudo for MVPA) were 40 for LST and 38 for MVPA.

Figure 3 shows the proportion of potential effects of higher LST and lower MVPA on diseases that passed the Bonferroni threshold, within broad disease categories (ICD-10 chapters). For all of these, higher LST and/or lower MVPA were found to contribute to increased disease risk. Most effects were observed in diseases of the musculoskeletal system and connective tissue (e.g., arthropathies, dorsopathies, soft tissue disorders), followed by diseases of the respiratory system (e.g., asthma, chronic lower respiratory diseases) and the genitourinary system (e.g., LSIL [Low-Grade Squamous Intraepithelial Lesion] lesion of the cervix uteri, vagina or vulva, excessive, frequent and irregular menstruation). However, no significant effects of either LST or lower MVPA were observed for certain infectious and parasitic diseases, diseases of the blood and blood-forming organs and certain disorders involving the immune mechanism, injury, poisoning and certain other consequences of external causes, external causes of morbidity and mortality or codes for special purposes.

**Figure 3.**
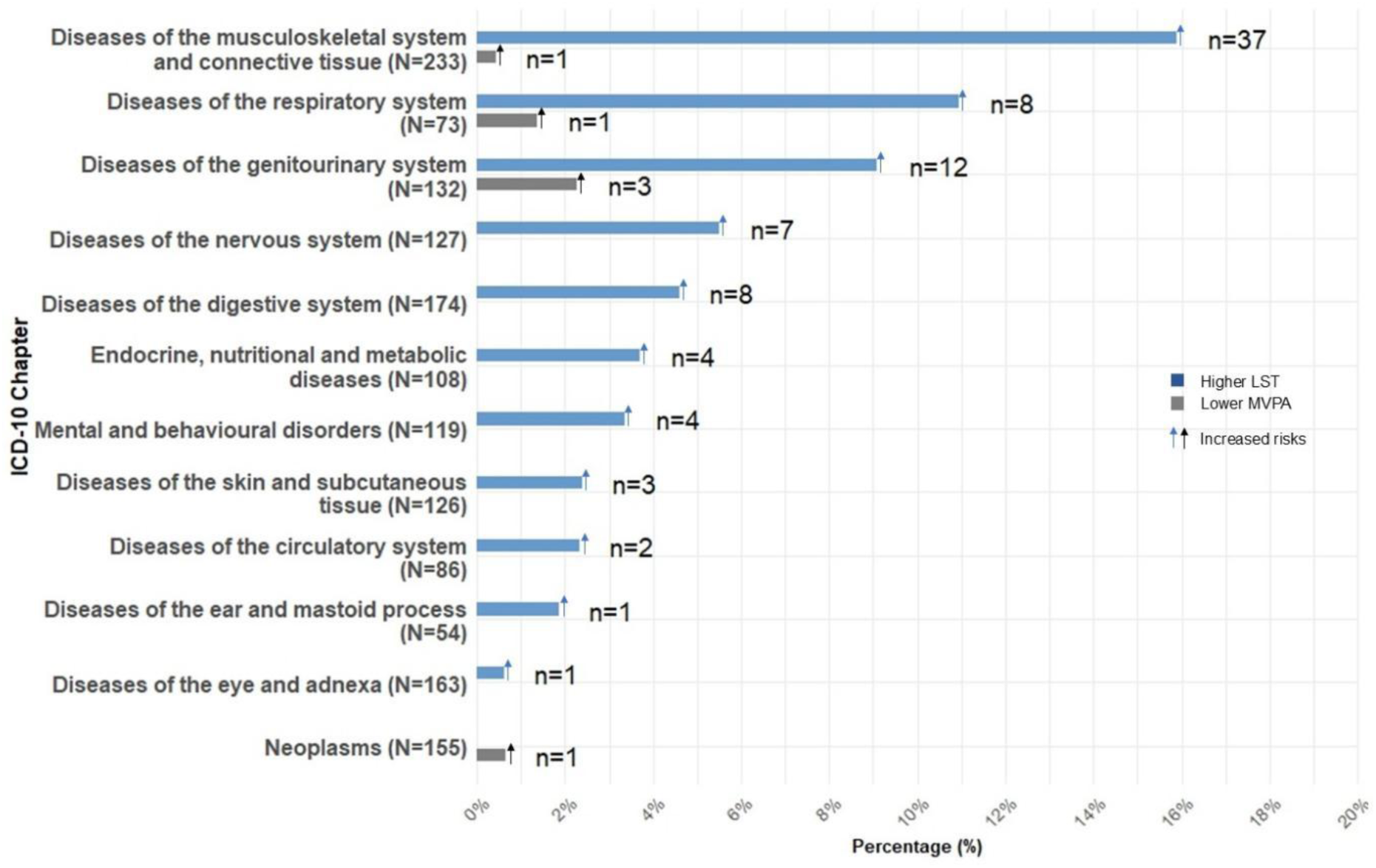
Percentage (and frequency) of diseases, within each ICD-10 chapter, showing significant effects of higher LST and/or lower MVPA based on Bonferroni correction: Notes: No significant effects were found for the following ICD-10 chapters, based on Bonferroni correction: certain infectious and parasitic diseases (N=97); diseases of the blood and blood-forming organs and certain disorders involving the immune mechanism (N=55); injury, poisoning and certain other consequences of external causes (N=14); external causes of morbidity and mortality (N=1) and codes for special purposes (N=2). Higher LST: longer leisure screen time (hours per day). Lower MVPA: engaging in moderate-to-vigorous intensity physical activity for less than 20 minutes per week or once or less per week. n=number of significant diseases identified using the Bonferroni correction. N=total number of diseases in the chapter.

Based on the Bonferroni corrected threshold, we observed that higher LST had a potential effect of increasing the risk of 87 (5.1%) of the 1,719 diseases explored. Of these 87, 76 (87.4%) had estimated effects in the same direction for genetic liability to lower MVPA, with three of these reaching the Bonferroni-corrected threshold (Supplementary Figure S1). In addition to these three overlapping diseases, genetic liability to lower MVPA was associated with increased risks (following Bonferroni-correction) of non-small cell lung cancer, later onset COPD and unspecified/other endometriosis (Figure 4). Supplementary Table S6 gives the main IVW results for the effect of each exposure on all diseases grouped by chapters, with 95% confidence intervals (CIs), ordered by p-value.

**Figure 4.**
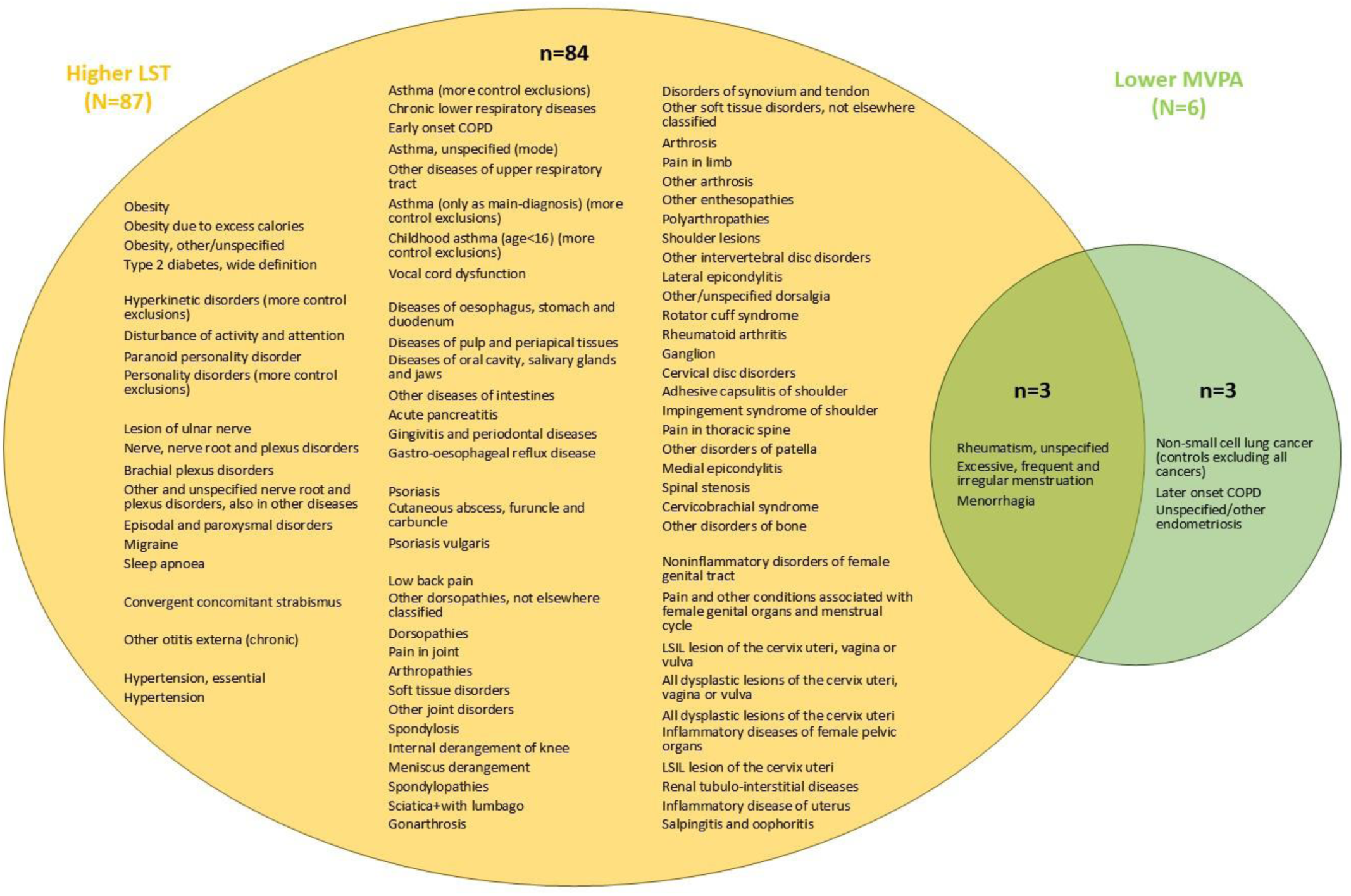
Shared and unique diseases identified as relating to higher LST and/or lower MVPA: Higher LST: higher leisure screen time (hours per day) Lower MVPA: engaging in moderate-to-vigorous intensity physical activity for less than 20 minutes per week or once or less per week. COPD: Chronic Obstructive Pulmonary Disease LSIL: Low-Grade Squamous Intraepithelial Lesion n=number of diseases related to higher LST or lower MVPA identified using the Bonferroni correction

### Sensitivity analyses

Results of the sensitivity analyses are presented in Supplementary Table S6. Based on Cochran’s Q statistic, evidence of SNP heterogeneity was observed for 67 of the 87 diseases affected by higher LST, and for two of the three diseases affected by lower MVPA.

The effects identified in the discovery stage were consistent across sensitivity analyses (Supplementary Figure S2). The weighted median method supported the effects of higher LST, and genetic liability to lower MVPA, on the diseases identified in the discovery analyses. Most diseases (82/87 for higher LST; 6/6 for lower MVPA) showed no evidence of horizontal pleiotropy and the majority of MR-Egger estimates (64/87 for higher LST; 5/6 for lower MVPA) were consistent in direction with those obtained in the discovery analyses. Both higher LST, and genetic liability to lower MVPA, showed consistent effects on diseases after applying MR Steiger filtering.

### Replication analyses

Of the 87 diseases with potential effects of LST, UK Biobank GWAS data were available for 65; 51 of these replicated with an effect in the same direction and p ≤ 0.05, 11 showed directional consistency only and three showed no directional consistency. For lower MVPA, GWAS data were available for four of the six diseases; one replicated, and the remaining three showed directional consistency. Detailed results are provided in Supplementary Table S5.

## Discussion

In this large hypothesis-free MR-PheWAS, we found evidence of potential causal effects of sedentary behaviour (LST), with higher LST increasing the risk of 87 (5.1%) of the 1,719 diseases explored, most of which were in musculoskeletal and connective tissue, respiratory and genitourinary systems. Although we had lower power to detect effects of lower levels of physical activity (MVPA), the results were largely what we would expect given the correlation of this exposure with LST. We observed potential causal effects of increased risk of diseases in musculoskeletal and connective tissue, genitourinary and respiratory systems that were consistent with LST findings. In addition, of the 87 diseases that were significant for LST, 76 had estimated effects in the same direction for genetic liability to lower MVPA. Sensitivity analyses found all of these potential effects supported by weighted median method and most by MR-Egger. We were able to find GWAS for most of these potential effects for replication in UK Biobank, and 51 of the 65 effects were replicated, showing directional consistency and p ≤ 0.05, with 11 of the remainder being directionally consistent but having imprecise estimates.

Adverse effects of greater sedentary behaviour on diseases of the musculoskeletal system and connective tissue (e.g., osteoarthritis, low back pain, sclerosis, etc.) and respiratory system (e.g., COPD, asthma, etc.) are consistent with findings from previous MR studies, whereas few such past studies have explored associations with genitourinary diseases (see Supplementary Table S1). Our finding of potential effects of LST and MVPA on increased risk of genitourinary diseases that have previously rarely been explored highlights the importance of hypothesis-free approaches that go beyond outcomes that are repeatedly studied. The diseases we identified as being potentially caused by high levels of sedentary behaviour and low physical activity, that were largely female reproductive conditions – such as cervix uteri, vagina or vulva conditions – suggest a possible role for physical activity in female pelvis strength, whereas the potential effect on menorrhagia may reflect the role of higher body mass index in polycystic ovary syndrome and reproductive hormones [39]. Conversely, it is possible that these findings reflect reverse causality (i.e. women with these conditions may be less able to exercise and thus more sedentary). However, Steiger filtering analyses suggested that these and other potential effects were unlikely due to reverse causality. Further research of how physical activity might affect women’s genitourinary / reproductive system is warranted.

We detected borderline evidence of a causal effect of high sedentary behaviour or low physical activity on an increased risk of depression. This is somewhat consistent with systematic reviews of randomised controlled trials (RCTs) suggesting that interventions to increase physical activity have little or only modest effect on depression [40–42].

Sedentary behaviour affected fewer cardiovascular diseases than reported in previous studies [43, 44], with the observed effects appearing to be primarily driven by hypertension, indicating that hypertension may be the cardiovascular disease most sensitive to sedentary behaviour. In relation to cardiovascular disease, RCTs of physical activity interventions have provided evidence that exercise-based cardiac rehabilitation reduces the risk of cardiovascular events [45, 46].

The smaller number of cardiovascular diseases detected in this study may reflect limited statistical power to identify true effects after applying our relatively conservative criteria. For example, we observed the same direction of effect (1.08 [1.01, 1.17]) of sedentary behaviour on ischaemic heart disease as reported in previous MR studies [43, 47]. Bonferroni and FDR corrections can reduce false positives, but they may also increase the likelihood of false negatives [48]; that is, true effects can be overlooked. However, because this study employs a hypothesis-free approach, a stringent threshold is necessary to limit false positives, whereas hypothesis-driven studies can use a less stringent threshold (e.g., p≤0.05).

There are several interventions focusing on encouraging physical activities and reducing sedentary behaviours to promote health-related outcomes. For example, an intervention to reduce sitting time among office workers showed small improvements in stress, wellbeing, vigour, and pain in the lower extremity [49]. The adverse effects of higher sedentary behaviour and lower physical activity across multiple systems observed in this study indicate that interventions aimed at reducing sedentary behaviour and increasing moderate-to-vigorous physical activity could be beneficial for preventing multimorbidity.

## Study strengths and limitations

This study had several strengths. First, we applied MR-PheWAS, a hypothesis-free causal inference approach, which enabled us to systematically investigate the effects of sedentary behaviour and physical activity on 1,719 diseases. This method helps elucidate any causal role of these exposures on multimorbidity and allows detection of effects which may not have been previously investigated. Second, diseases were defined using national health registries in FinnGen, which capture patients with both mild and severe conditions, thereby minimising misclassification bias. Third, the analyses were conducted in a large sample, and stringent criteria for effect detection were applied, reducing the chance of false positives. Finally, we performed sensitivity analyses to address potential violations of core MR assumptions and conducted replication analyses in UK Biobank with available GWAS. The high consistency of these results further strengthened the robustness and reliability of our findings.

This study also has limitations. Despite reducing false positives, Bonferroni correction increases false negatives, which may lead to missing some true effects of our exposures. Given MVPA is a binary exposure here, it is likely we have lower statistical power to detect its effects compared to LST, a continuous exposure [20]. Thus, estimates indicating possible effects of lower MVPA on disease that did not pass Bonferroni correction may be considered suggestive, although future replication in higher-powered analyses is needed. Measurements of LST and MVPA as used in the GWAS meta-analyses were based on self-reported items; for MVPA, in particular, these often differed from study-to-study, which may lead to measurement error and invalid genetic instruments, although it is somewhat reassuring that modest genetic correlations have been observed between self-reported and accelerometer-assessed physical activity traits [21]. Whilst it is important to adjust for as many markers of population substructure as possible for the MR independence assumption, in our two-sample MR we were limited to the adjustments for ancestral PCs – which may be fewer than optimal – made in the original GWAS. The exposure and outcome GWAS in the replication analyses showed considerable population overlap, which may bias estimates away from the null, although any effect is likely small [50]. We excluded diseases with small case numbers (<100) which means we were not able to detect effects on rare diseases. We were unable to test whether all effects identified in the discovery phase of our MR-PheWAS were replicated, because not all ICD-10-coded diseases available in FinnGen were represented in UK Biobank. This study focused only on participants of European ancestry, which limits the generalizability of the findings to other populations with different genetic backgrounds. Studies based on larger sample sizes and non-European populations or trans-ancestry analyses are needed [51].

## Conclusions

Higher levels of sedentary behaviour, and lower levels of moderate-to-vigorous physical activity, increase the risk of diseases across multiple systems, primarily the musculoskeletal system and connective tissue and genitourinary and respiratory systems. While some of these effects were previously established, most identified in this study are novel, including those related to genitourinary diseases. These findings suggest that reducing sedentary behaviour, and the corollary of increasing moderate-to-vigorous physical activity, will help lower the risk of multimorbidity.

## Supporting information

### S1 Appendix

**Supplementary Material S1.** GWAS for sedentary behaviour and physical activity. **Supplementary Figure S1.** Forest plot of diseases with estimated causal effects of LST or lower MVPA passing the Bonferroni-corrected p-value threshold, grouped by ICD-10 blocks. **Supplementary Figure S2.** Sensitivity analyses of diseases with LST effects passing the Bonferroni-corrected p-value threshold, grouped by ICD-10 chapters.

### S2 Appendix

**Supplementary Table S1-1.** Studies investigating causal effects of physical activity or sedentary behaviour on diseases using Mendelian randomization (MR). **Supplementary Table S1-2.** Summary of studies investigating causal effects of physical activity or sedentary behaviour on diseases using Mendelian randomization (MR). **Supplementary Table S2.** Characteristics of contributing studies of European ancestry and potential sample overlap. **Supplementary Table S3.** Instrumental variables based on 5*10^-8. **Supplementary Table S4.** Inclusion and exclusion for FinnGen R11 endpoints. **Supplementary Table S5.** Replication analyses in the UK Biobank for diseases that passed Bonferroni correction in MR-PheWAS. **Supplementary Table S6-1.** MR-PheWAS results for certain infectious and parasitic diseases. **Supplementary Table S6-2.** MR-PheWAS results for neoplasms. **Supplementary Table S6-3.** MR-PheWAS results for diseases of the blood and blood-forming organs and certain disorders involving the immune mechanism. **Supplementary Table S6-4.** MR-PheWAS results for endocrine, nutritional and metabolic diseases. **Supplementary Table S6-5.** MR-PheWAS results for mental and behavioural disorders. **Supplementary Table S6-6.** MR-PheWAS results for diseases of the nervous system. **Supplementary Table S6-7.** MR-PheWAS results for diseases of the eye and adnexa. **Supplementary Table S6-8.** MR-PheWAS results for diseases of the ear and mastoid process. **Supplementary Table S6-9.** MR-PheWAS results for diseases of the circulatory system. **Supplementary Table S6-10.** MR-PheWAS results for diseases of the respiratory system. **Supplementary Table S6-11.** MR-PheWAS results for diseases of the digestive system. **Supplementary Table S6-12.** MR-PheWAS results for diseases of the skin and subcutaneous tissue. **Supplementary Table S6-13.** MR-PheWAS results for diseases of the musculoskeletal system and connective tissue. **Supplementary Table S6-14.** MR-PheWAS results for diseases of the genitourinary system. **Supplementary Table S6-15.** MR-PheWAS results for injury, poisoning and certain other consequences of external causes. **Supplementary Table S6-16.** MR-PheWAS results for external causes of morbidity and mortality. **Supplementary Table S6-17.** MR-PheWAS results for codes for special purposes.

## S3 STROBE-MR-checklist

### Availability of data and materials

FinnGen genetic summary statistics are available online (https://finngen.gitbook.io/documentation/r11/data-download). UK Biobank genetic summary data can be downloaded from the Neale lab (https://www.nealelab.is/uk-biobank), GeneATLAS (http://geneatlas.roslin.ed.ac.uk/downloads/) and OpenGWAS platform (https://gwas.mrcieu.ac.uk/datasets). Analysis plan and code for this study are available on Github (https://github.com/JiayaoXu2023/MR-PheWAS-SB-PA).

## Author Contributions

DAL, GLC, JX, RMAP, KB conceived and designed the study and wrote the analysis plan. JX conducted the data analysis and wrote the original draft of the manuscript, with reviewing and editing by DAL, GLC, RMAP, KB.

## Supporting information

S3 STROBE-MR-checklist

## Acknowledgements

We want to acknowledge the participants and investigators of the FinnGen study. We are grateful to the UK Biobank participants for making this research possible.

