## Supplementary material for "The effect of sedentary behaviour and physical activity on 1719 diseases: a Mendelian randomisation phenome-wide association study (MR-PheWAS)": S1 Appendix

### Supplementary Material S1 GWAS for sedentary behaviour and physical activity

In the GWAS paper on LST and MVPA, the authors conducted separate GWAS analyses within each study, stratified by sex and ancestry [1]. Additive genetic models, accounting for family relatedness when appropriate, were adjusted for age, age-squared, principal components representing population structure and additional study-specific covariates (e.g., genotyping platform, genotyping calling method, imputation methods). Family relatedness was accounted for in the models where appropriate. Analyses were conducted on genotyped and imputed variants with a minor allele frequency > 0.1% in UK Biobank and a minor allele count >3 in other studies. GWAS results were meta-analysed in METAL [2] using a fixed-effects, inverse variance-weighted approach. In the original GWAS paper [1], the authors used a GWAS significance threshold of $p < 5\times10^{-9}$ to select SNPs, as they did not have a replication stage. The GWAS indicated modest genetic correlations between accelerometer-assessed physical activity traits and both self-reported MVPA and LST.


1. Wang, Z., et al., *Genome-wide association analyses of physical activity and sedentary behavior provide insights into underlying mechanisms and roles in disease prevention*. Nature Genetics, 2022. **54**(9): p. 1332-+.
2. Willer, C.J., Y. Li, and G.R. Abecasis, *METAL: fast and efficient meta-analysis of genomewide association scans*. Bioinformatics, 2010. **26**(17): p. 2190-1.

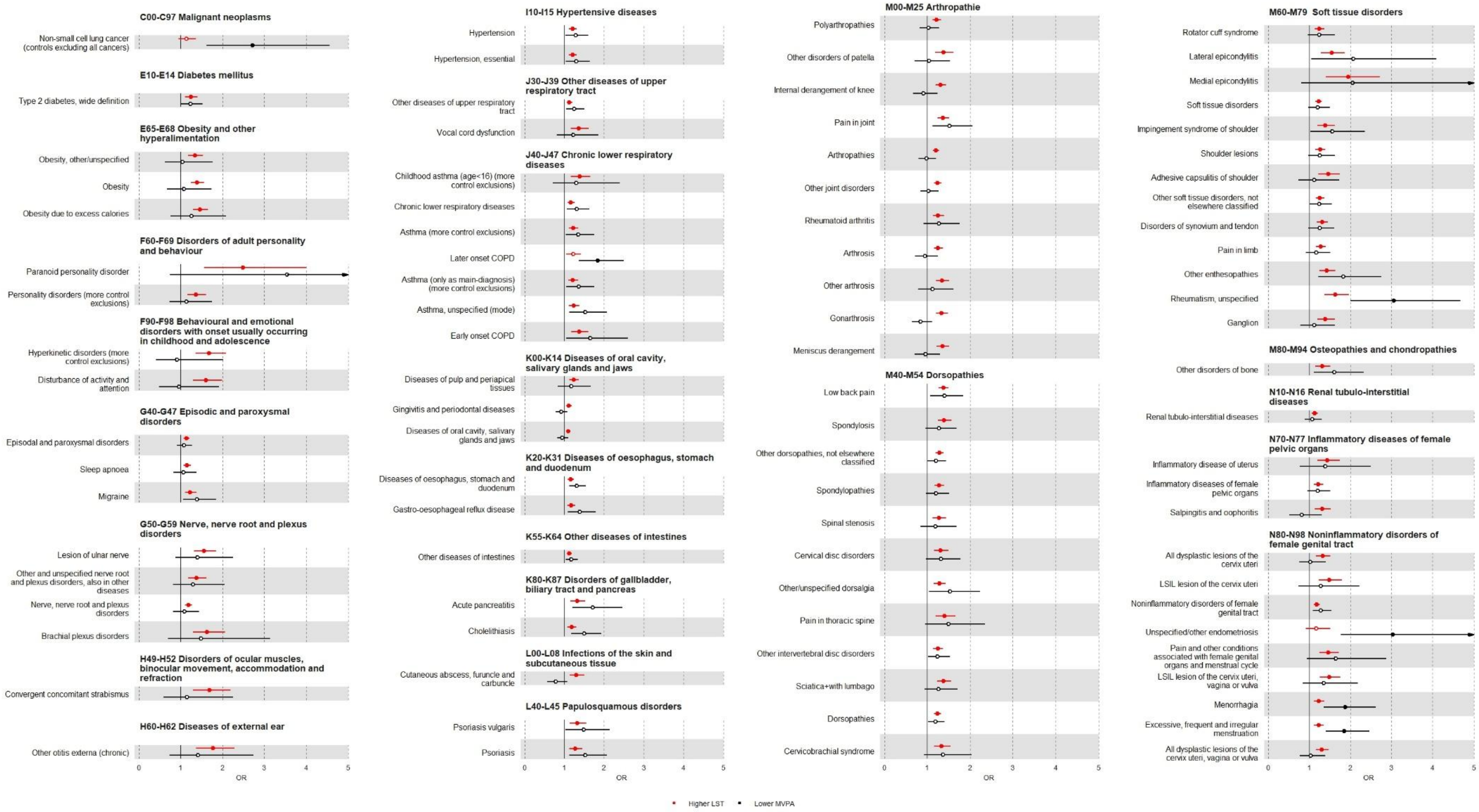
C00-C97 Malignant neoplasms
Non-small cell lung cancer (controls excluding all cancers)
E10-E14 Diabetes mellitus
Type 2 diabetes, wide definition
E65-E68 Obesity and other hyperalimentation
Obesity, other/unspecified
Obesity
Obesity due to excess calories
F60-F69 Disorders of adult personality and behaviour
Paranoid personality disorder
Personality disorders (more control exclusions)
F90-F98 Behavioural and emotional disorders with onset usually occurring in childhood and adolescence
Hyperkinetic disorders (more control exclusions)
Disturbance of activity and attention
G40-G47 Episodic and paroxysmal disorders
Episodal and paroxysmal disorders
Sleep apnoea
Migraine
G50-G59 Nerve, nerve root and plexus disorders
Lesion of ulnar nerve
Other and unspecified nerve root and plexus disorders, also in other diseases
Nerve, nerve root and plexus disorders
Brachial plexus disorders
H49-H52 Disorders of ocular muscles, binocular movement, accommodation and refraction
Convergent concomitant strabismus
H60-H62 Diseases of external ear
Other otitis externa (chronic)
0
1
2
3
4
5
OR
I10-I15 Hypertensive diseases
Hypertension
Hypertension, essential
J30-J39 Other diseases of upper respiratory tract
Other diseases of upper respiratory tract
Vocal cord dysfunction
J40-J47 Chronic lower respiratory diseases
Childhood asthma (age<16) (more control exclusions)
Chronic lower respiratory diseases
Asthma (more control exclusions)
Later onset COPD
Asthma (only as main-diagnosis) (more control exclusions)
Asthma, unspecified (mode)
Early onset COPD
K00-K14 Diseases of oral cavity, salivary glands and jaws
Diseases of pulp and periapical tissues
Gingivitis and periodontal diseases
Diseases of oral cavity, salivary glands and jaws
K20-K31 Diseases of oesophagus, stomach and duodenum
Diseases of oesophagus, stomach and duodenum
Gastro-oesophageal reflux disease
K55-K64 Other diseases of intestines
Other diseases of intestines
K80-K87 Disorders of gallbladder, biliary tract and pancreas
Acute pancreatitis
Cholelithiasis
L00-L08 Infections of the skin and subcutaneous tissue
Cutaneous abscess, furuncle and carbuncle
L40-L45 Papulosquamous disorders
Psoriasis vulgaris
Psoriasis
0
1
2
3
4
5
OR
M00-M25 Arthropathie
Polyarthropathies
Other disorders of patella
Internal derangement of knee
Pain in joint
Arthropathies
Other joint disorders
Rheumatoid arthritis
Arthrosis
Other arthrosis
Gonarthrosis
Meniscus derangement
M40-M54 Dorsopathies
Low back pain
Spondylosis
Other dorsopathies, not elsewhere classified
Spondylopathies
Spinal stenosis
Cervical disc disorders
Other/unspecified dorsalgia
Pain in thoracic spine
Other intervertebral disc disorders
Sciatica+with lumbago
Dorsopathies
Cervicobrachial syndrome
0
1
2
3
4
5
OR
M60-M79 Soft tissue disorders
Rotator cuff syndrome
Lateral epicondylitis
Medial epicondylitis
Soft tissue disorders
Impingement syndrome of shoulder
Shoulder lesions
Adhesive capsulitis of shoulder
Other soft tissue disorders, not elsewhere classified
Disorders of synovium and tendon
Pain in limb
Other enthesopathies
Rheumatism, unspecified
Ganglion
M80-M94 Osteopathies and chondropathies
Other disorders of bone
N10-N16 Renal tubulo-interstitial diseases
Renal tubulo-interstitial diseases
N70-N77 Inflammatory diseases of female pelvic organs
Inflammatory disease of uterus
Inflammatory diseases of female pelvic organs
Salpingitis and oophoritis
N80-N98 Noninflammatory disorders of female genital tract
All dysplastic lesions of the cervix uteri
LSIL lesion of the cervix uteri
Noninflammatory disorders of female genital tract
Unspecified/other endometriosis
Pain and other conditions associated with female genital organs and menstrual cycle
LSIL lesion of the cervix uteri, vagina or vulva
Menorrhagia
Excessive, frequent and irregular menstruation
All dysplastic lesions of the cervix uteri, vagina or vulva
0
1
2
3
4
5
OR
Higher LST
Lower MVPA

Notes: LST: leisure screen time, MVPA: moderate-to-vigorous intensity physical activity; OR for LST: odds ratio of each disease per one-hour increase in leisure screen time (hours per day); OR for lower MVPA compared to higher MVPA: odds ratio of each disease per unit increase in the genetically predicted log-odds of engaging in less than 20 minutes of MVPA per week (compared to ≥20 minutes) or once or less per week (compared to twice or more per week). Solid dots indicate statistically significant estimates based on the Bonferroni corrected threshold ($p \leq 3.47 \times 10^{-4}$)

**Supplementary Figure S1 Forest plot of diseases with estimated causal effects of LST or lower MVPA passing the Bonferroni-corrected p-value threshold, grouped by ICD-10 blocks**

a Diseases of the musculoskeletal system and connective tissue

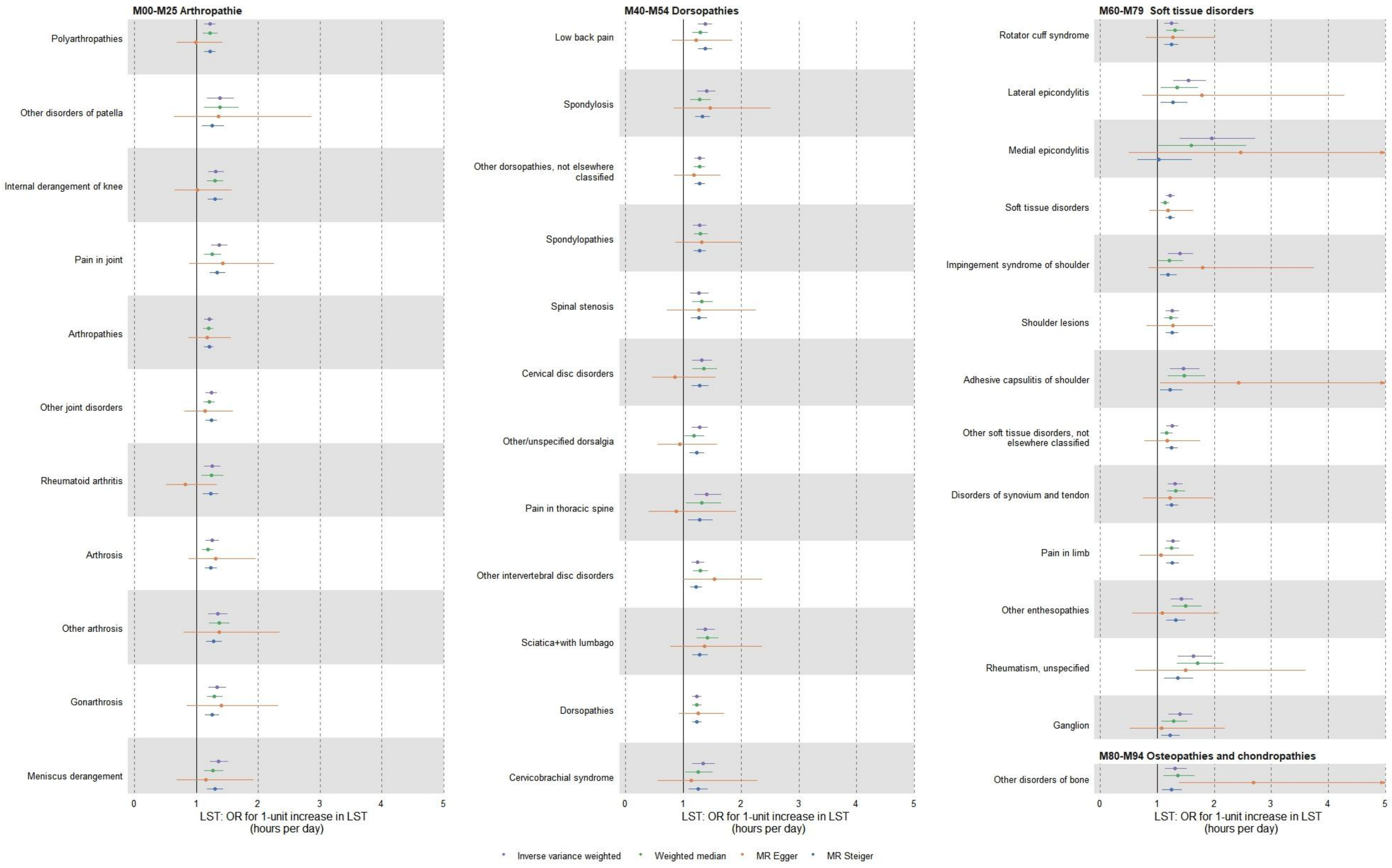

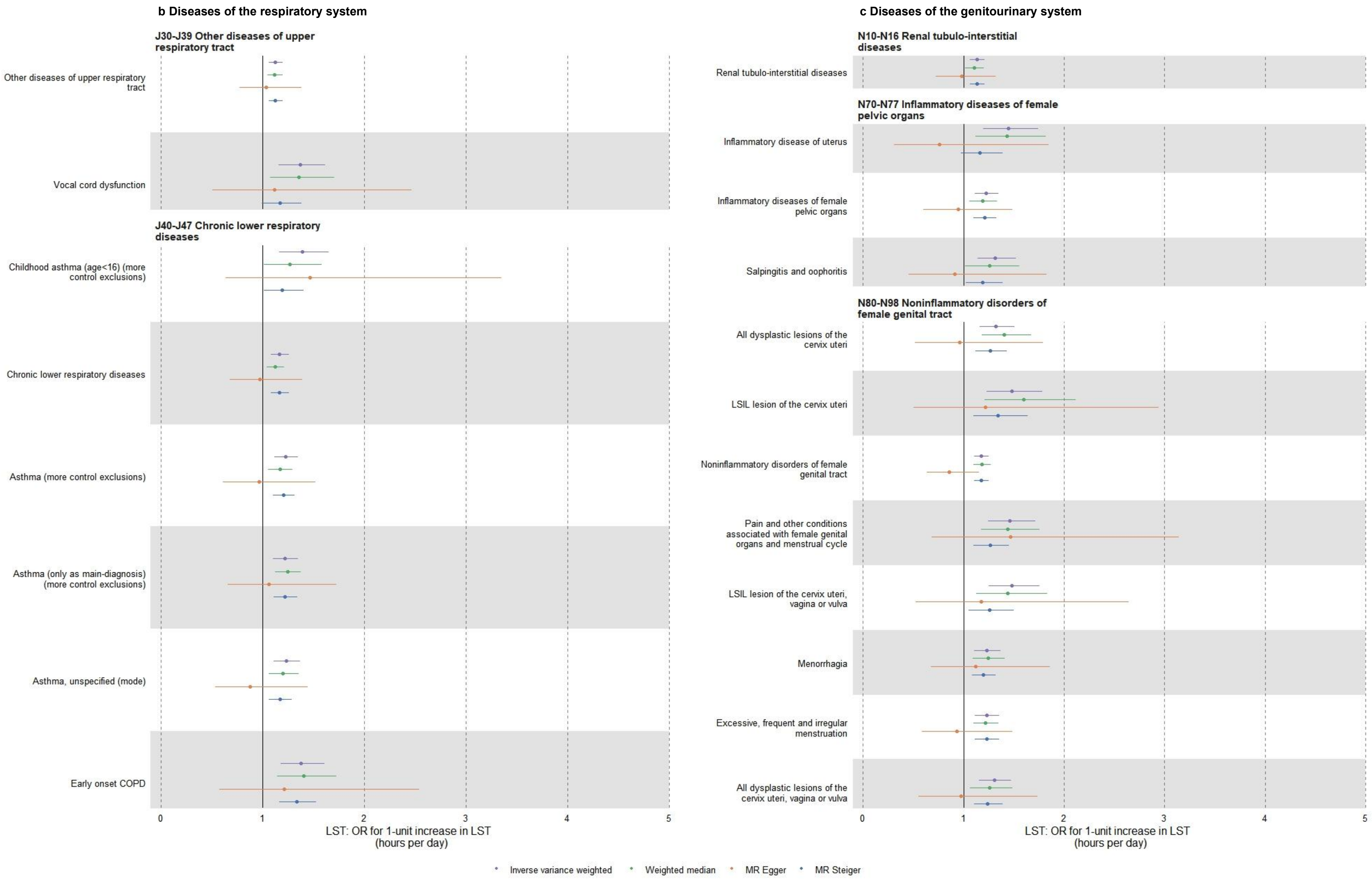

b Diseases of the respiratory system
J30-J39 Other diseases of upper respiratory tract
Other diseases of upper respiratory tract
Vocal cord dysfunction
J40-J47 Chronic lower respiratory diseases
Childhood asthma (age<16) (more control exclusions)
Chronic lower respiratory diseases
Asthma (more control exclusions)
Asthma (only as main-diagnosis) (more control exclusions)
Asthma, unspecified (mode)
Early onset COPD
0
1
2
3
4
5
LST: OR for 1-unit increase in LST (hours per day)
c Diseases of the genitourinary system
N10-N16 Renal tubulo-interstitial diseases
Renal tubulo-interstitial diseases
N70-N77 Inflammatory diseases of female pelvic organs
Inflammatory disease of uterus
Inflammatory diseases of female pelvic organs
Salpingitis and oophoritis
N80-N98 Noninflammatory disorders of female genital tract
All dysplastic lesions of the cervix uteri
LSIL lesion of the cervix uteri
Noninflammatory disorders of female genital tract
Pain and other conditions associated with female genital organs and menstrual cycle
LSIL lesion of the cervix uteri, vagina or vulva
Menorrhagia
Excessive, frequent and irregular menstruation
All dysplastic lesions of the cervix uteri, vagina or vulva
0
1
2
3
4
5
LST: OR for 1-unit increase in LST (hours per day)
Inverse variance weighted
Weighted median
MR Egger
MR Steiger

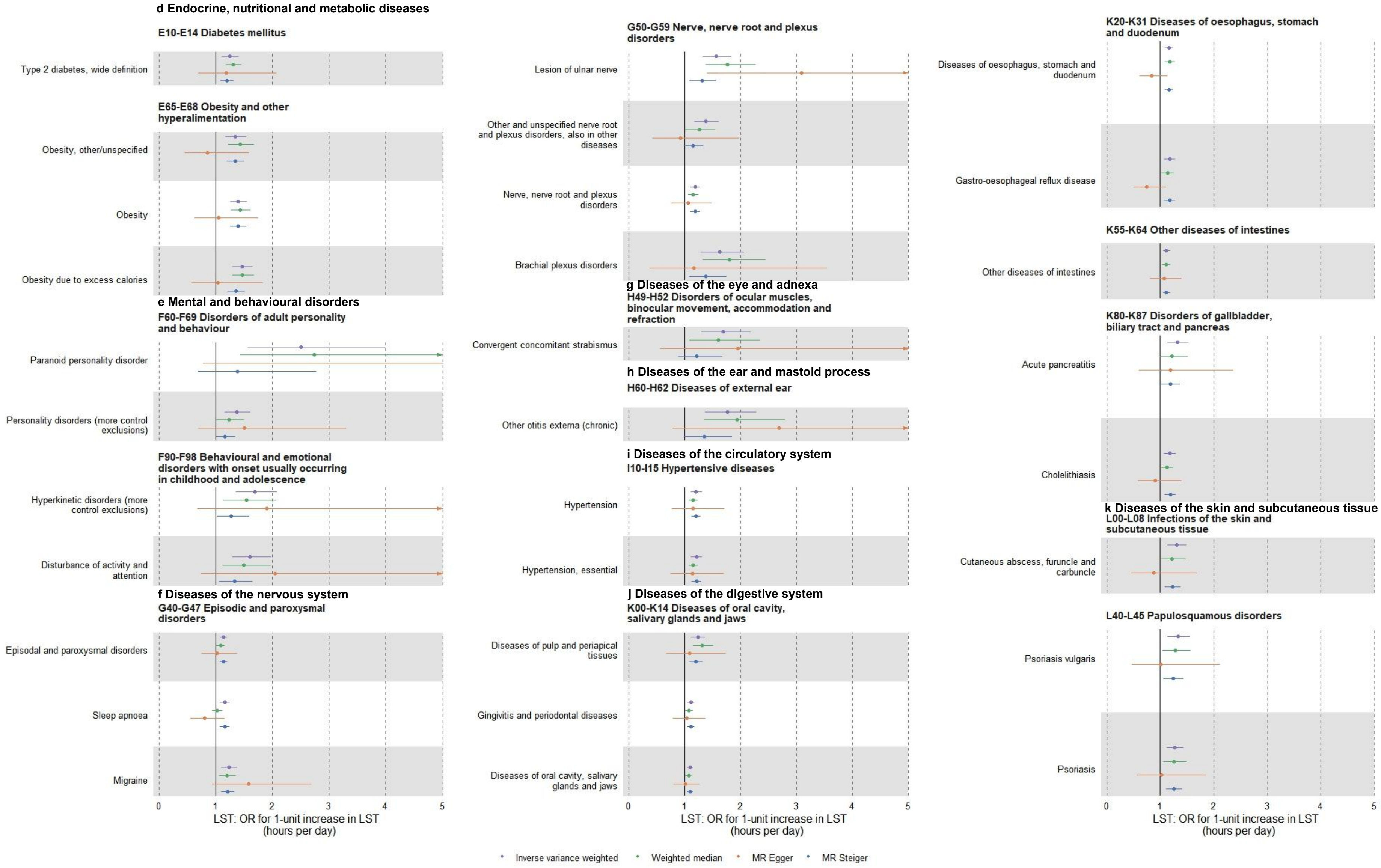
d Endocrine, nutritional and metabolic diseases
E10-E14 Diabetes mellitus
Type 2 diabetes, wide definition
E65-E68 Obesity and other hyperalimentation
Obesity, other/unspecified
Obesity
Obesity due to excess calories
e Mental and behavioural disorders
F60-F69 Disorders of adult personality and behaviour
Paranoid personality disorder
Personality disorders (more control exclusions)
F90-F98 Behavioural and emotional disorders with onset usually occurring in childhood and adolescence
Hyperkinetic disorders (more control exclusions)
Disturbance of activity and attention
f Diseases of the nervous system
G40-G47 Episodic and paroxysmal disorders
Episodal and paroxysmal disorders
Sleep apnoea
Migraine
G50-G59 Nerve, nerve root and plexus disorders
Lesion of ulnar nerve
Other and unspecified nerve root and plexus disorders, also in other diseases
Nerve, nerve root and plexus disorders
Brachial plexus disorders
g Diseases of the eye and adnexa
H49-H52 Disorders of ocular muscles, binocular movement, accommodation and refraction
Convergent concomitant strabismus
h Diseases of the ear and mastoid process
H60-H62 Diseases of external ear
Other otitis externa (chronic)
i Diseases of the circulatory system
I10-I15 Hypertensive diseases
Hypertension
Hypertension, essential
j Diseases of the digestive system
K00-K14 Diseases of oral cavity, salivary glands and jaws
Diseases of pulp and periapical tissues
Gingivitis and periodontal diseases
Diseases of oral cavity, salivary glands and jaws
K20-K31 Diseases of oesophagus, stomach and duodenum
Diseases of oesophagus, stomach and duodenum
Gastro-oesophageal reflux disease
K55-K64 Other diseases of intestines
Other diseases of intestines
K80-K87 Disorders of gallbladder, biliary tract and pancreas
Acute pancreatitis
Cholelithiasis
k Diseases of the skin and subcutaneous tissue
L00-L08 Infections of the skin and subcutaneous tissue
Cutaneous abscess, furuncle and carbuncle
L40-L45 Papulosquamous disorders
Psoriasis vulgaris
Psoriasis
0
1
2
3
4
5
LST: OR for 1-unit increase in LST (hours per day)
Inverse variance weighted
Weighted median
MR Egger
MR Steiger

Notes: LST: leisure screen time (hours per day); Inverse variance weighted: inverse-variance weighted method; Weighted median: weighted median method; MR Egger: MR-Egger method; MR Steiger: Steiger filtering

**Supplementary Figure S2 Sensitivity analyses of diseases with LST effects passing the Bonferroni-corrected p-value threshold, grouped by ICD-10 chapters**
