## Supplementary material for "The effect of sedentary behaviour and physical activity on 1719 diseases: a Mendelian randomisation phenome-wide association study (MR-PheWAS)": S3 STROBE-MR-checklist

### STROBE-MR checklist of recommended items to address in reports of Mendelian randomization studies[1 2]

| Item No. | Section | | Checklist item | Page No. | Relevant text from manuscript |
|---|---|---|---|---|---|
| 1 | **TITLE and ABSTRACT** | | Indicate Mendelian randomization (MR) as the study's design in the title and/or the abstract if that is a main purpose of the study | 1 | The effect of sedentary behaviour and physical activity on 1719 diseases: a Mendelian randomisation phenome-wide association study (MR-PheWAS) |
| | **INTRODUCTION** | | | | |
| 2 | **Background** | | Explain the scientific background and rationale for the reported study. What is the exposure? Is a potential causal relationship between exposure and outcome plausible? Justify why MR is a helpful method to address the study question | 5 | Mendelian randomization (MR), which typically uses genetic variants as instrumental variables for exposures, is widely used to explore unconfounded effects of exposures on outcomes in observational data, under specific assumptions [11]. As genetic variants are randomly assorted at conception, they are less likely to be affected by confounding and reverse causation than conventional multivariable regression [12]. MR phenome-wide association studies (MR-PheWAS) are an extension of MR that test the effects of one or more exposures on a wide range of diseases, selected in a hypothesis-free way [13]. |
| 3 | **Objectives** | | State specific objectives clearly, including pre-specified causal hypotheses (if any). State that MR is a method that, under specific assumptions, intends to estimate causal effects | 5-6 | Mendelian randomization (MR), which typically uses genetic variants as instrumental variables for exposures, is widely used to explore unconfounded effects of exposures on outcomes in observational data, under specific assumptions [11].<br>The primary aim of this study was to explore the effects of higher sedentary behaviour and, secondarily, lower physical activity on diseases across multiple systems. |
| | **METHODS** | | | | |
| 4 | **Study design and data sources** | | Present key elements of the study design early in the article. Consider including a table listing sources of data for all phases of the study. For each data source contributing to the analysis, describe the following: | | |
| | | a) | Setting: Describe the study design and the underlying population, if possible. Describe the setting, locations, and relevant dates, including periods of recruitment, exposure, follow-up, and data collection, when available. | 6 | Figure 1 summarises the steps in our MR PheWAS. First, we identified independent genetic variants to instrument LST and moderate-to-vigorous intensity physical activity (MVPA). We then explored the associations of those single |

| | | | |
|---|---|---|---|
| | | | nucleotide polymorphisms (SNPs) with disease outcomes in the discovery data (FinnGen [17]), ensuring that the alleles were harmonised to match those used as the genetic instruments. For main MR analyses, we used inverse variance weighted (IVW) method, with 1,719 diseases included within broad ICD-10 chapters. Whilst these diseases are distinct, they are not all independent within chapters, so a Bonferroni corrected p-value of $p \leq 0.05/144$ ($3.47\times10^{-4}$) was used based on the total number of ICD-10 blocks (n=144), the tier of disease classification below that of chapter. Effects for LST and MVPA that passed the threshold were taken forward for MR sensitivity analyses to test bias, and also for replication in UK Biobank [18], for which we repeated the main IVW analyses for diseases with available GWAS data. Results were defined as replicating if the direction of effect in UK Biobank was the same as that in the FinnGen discovery analyses, and $p \leq 0.05$. |
| b) | Participants: Give the eligibility criteria, and the sources and methods of selection of participants. Report the sample size, and whether any power or sample size calculations were carried out prior to the main analysis | 9-12 | This GWAS meta-analysed results from 51 studies comprising 661,399 European adults, with UK Biobank contributing 68% of the sample [21] (Supplementary Table S2).<br><br>It includes a nationally representative adult Finnish population with a median age of 63 years [24]. We used the most recent data release (R11) of disease GWAS summary results available when starting this study (August 2024). This included 453,733 participants of European ancestry.<br><br>UK Biobank is a UK prospective cohort study that enrolled 503,325 adults (5.5% of those invited) aged 40 to 69, with 91% identifying as white European [18]. Participants were recruited across 22 assessment centres between 2006 and 2010. |
| c) | Describe measurement, quality control and selection of genetic variants | 9-10 | In this GWAS, LST was analysed as a continuous measure of hours of television watched per day (24 hours). MVPA was analysed as a dichotomous exposure, with "higher MVPA" defined as ≥20 minutes versus <20 minutes per week for studies reporting duration (n=35), and ≥2 versus <2 times per week for studies reporting frequency (n=3) [13]. In all studies contributing to the GWAS, both exposures were assessed by self-report, with the |

| | | | | | |
|---|---|---|---|---|---|
| | | | | | specific questionnaires listed in Supplementary Table S2.<br>To be consistent with LST, we coded “higher MVPA” as 0 (reference category) and “lower MVPA” as 1. We initially selected 7,462 SNPs for LST and 1,374 SNPs for MVPA based on genome-wide significance threshold ($p < 5×10-8$). To ensure that only independent SNPs were included as genetic instrumental variables, we identified correlated SNPs defined as linkage disequilibrium (LD) $r2 >0.001$, within a 10,000 kb window using the 1000 Genomes European reference panel [22]. Where we found groups of correlated SNPs, we selected the one with the lowest p-value. Following this, we retained 117 independent SNPs for the LST genetic instrument and 18 independent SNPs for the lower MVPA genetic instrument (Supplementary Table S3). |
| | | d) | For each exposure, outcome, and other relevant variables, describe methods of assessment and diagnostic criteria for diseases | 10, 12 | From FinnGen’s list of clinical endpoints, we selected diseases identifiable in the hospital discharge registry that could be mapped to the International Classification of Diseases, Tenth Revision (ICD-10) codes, which is used in UK Biobank and therefore offered the potential to replicate findings in an independent large study. In FinnGen (discovery study), disease ICD codes were initially identified in the hospital discharge registry, and other registries were used only when an eligible disease was also recorded.<br>To explore replication of our FinnGen results, we used GWAS data for HES ICD-10 coded diseases in the UK Biobank |
| | | e) | Provide details of ethics committee approval and participant informed consent, if relevant | n/a | |
| 5 | **Assumptions** | | Explicitly state the three core IV assumptions for the main analysis (relevance, independence and exclusion restriction) as well assumptions for any additional or sensitivity analysis | 14-15 | MR has three core assumptions: 1) the genetic instruments are statistically strongly associated with the exposure in the relevant population (relevance assumption); 2) there is no confounding between the genetic instruments and disease (independence assumption); 3) the genetic instruments influence the disease(s) only through the exposure and not through any other pathway (exclusion restriction assumption) [35]. |

| | | | | | |
|---|---|---|---|---|---|
| | | | | | FinnGen across the whole genome-wide data irrespective of p-values. Where an exposure SNP was not present in the outcome GWAS, we explored whether a proxy SNP — defined as another SNP that is highly correlated with the target SNP (LD r2>0.8) — could be identified. We then harmonised the outcome SNPs to ensure consistency with the allele order of the corresponding exposure SNPs and checked that the harmonisation was successful. |
| | | c) | Describe the MR estimator (e.g. two-stage least squares, Wald ratio) and related statistics. Detail the included covariates and, in case of two-sample MR, whether the same covariate set was used for adjustment in the two samples | 9, 10, 13 | We used the inverse-variance weighted (IVW) method for the main MR analyses. The IVW method estimates the effect of interest using an inverse variance weighted regression of SNP-outcome associations on SNP-exposure associations, with the regression intercept constrained to zero [32].<br><br>Associations were adjusted for age, age2 and the top 10 ancestral principal components (PCs) (Supplementary Material S1).<br><br>The GWAS was adjusted for sex, age, 10 ancestry PCs, FinnGen chip version and legacy genotyping batch. |
| | | d) | Explain how missing data were addressed | 13 | For LST, 14 SNPs could not be harmonised because no suitable proxy was available for 10 missing SNPs and 4 SNPs were palindromic, resulting in 103 independent SNPs included in the genetic instrument. For MVPA, it was not possible to harmonise two SNPs because no suitable proxy was available for the missing SNPs; this resulted in 16 independent SNPs being included in the genetic instrument. |
| | | e) | If applicable, indicate how multiple testing was addressed | 6 | Whilst these diseases are distinct, they are not all independent within chapters, so a Bonferroni corrected p-value of p ≤ 0.05/144 (3.47×10-4) was used based on the total number of ICD-10 blocks (n=144), the tier of disease classification below that of chapter. |
| 7 | **Assessment of assumptions** | | Describe any methods or prior knowledge used to assess the assumptions or justify their validity | 14-15 | To assess these assumptions, we explored between-SNP heterogeneity tests for each exposure-outcome IVW analysis, using Cochran’s Q statistic. Between-SNP heterogeneity can occur when any of the assumptions are violated. We |

| | | | | | |
|---|---|---|---|---|---|
| 9 | **Software and pre-registration** | | | | |
| | | a) | Name statistical software and package(s), including version and settings used | 13 | The TwoSampleMR (MR-base) R package v 0.6.4 was used to extract and harmonise outcome SNPs and to run the analyses [31]. |
| | | b) | State whether the study protocol and details were pre-registered (as well as when and where) | 23 | Analysis plan and code for this study are available on Github (https://github.com/JiayaoXu2023/MR-PheWAS-SB-PA). |
| | **RESULTS** | | | | |
| 10 | **Descriptive data** | | | | |
| | | a) | Report the numbers of individuals at each stage of included studies and reasons for exclusion. Consider use of a flow diagram | 8 | Figure 1 Summary of study design |
| | | b) | Report summary statistics for phenotypic exposure(s), outcome(s), and other relevant variables (e.g. means, SDs, proportions) | 15 | We included 1,719 diseases in this MR-PheWAS. The mean F-statistics (pseudo for MVPA) were 40 for LST and 38 for MVPA. |
| | | c) | If the data sources include meta-analyses of previous studies, provide the assessments of heterogeneity across these studies | 9 | This GWAS meta-analysed results from 51 studies comprising 661,399 European adults, with UK Biobank contributing 68% of the sample [21] (Supplementary Table S2). |
| | | d) | For two-sample MR:<br>i. Provide justification of the similarity of the genetic variant-exposure associations between the exposure and outcome samples<br>ii. Provide information on the number of individuals who overlap between the exposure and outcome studies | 9, 10 | This GWAS meta-analysed results from 51 studies comprising 661,399 European adults, with UK Biobank contributing 68% of the sample [21] (Supplementary Table S2).<br>It includes a nationally representative adult Finnish population with a median age of 63 years [24]. We used the most recent data release (R11) of disease GWAS summary results available when starting this study (August 2024). This included 453,733 participants of European ancestry. |
| 11 | **Main results** | | | | |
| | | a) | Report the associations between genetic variant and exposure, and between genetic variant and outcome, preferably on an interpretable scale | 9-10 | Following this, we retained 117 independent SNPs for the LST genetic instrument and 18 independent SNPs for the lower MVPA genetic instrument (Supplementary Table S3). |

| | | | | | |
|---|---|---|---|---|---|
| | | b) | Report MR estimates of the relationship between exposure and outcome, and the measures of uncertainty from the MR analysis, on an interpretable scale, such as odds ratio or relative risk per SD difference | 17 | Supplementary Table S6 gives the main IVW results for the effect of each exposure on all diseases grouped by chapters, with 95% confidence intervals (CIs), ordered by p-value. |
| | | c) | If relevant, consider translating estimates of relative risk into absolute risk for a meaningful time period | n/a | |
| | | d) | Consider plots to visualize results (e.g. forest plot, scatterplot of associations between genetic variants and outcome versus between genetic variants and exposure) | 16 | Figure 3 Percentage (and frequency) of diseases, within each ICD-10 chapter, showing significant effects of higher LST and/or lower MVPA based on Bonferroni correction<br>Supplementary Figure S1 |
| 12 | **Assessment of assumptions** | | | | |
| | | a) | Report the assessment of the validity of the assumptions | 18 | Most diseases (82/87 for higher LST; 6/6 for lower MVPA) showed no evidence of horizontal pleiotropy |
| | | b) | Report any additional statistics (e.g., assessments of heterogeneity across genetic variants, such as $I^2$, Q statistic or E-value) | 18 | Based on Cochran's Q statistic, evidence of SNP heterogeneity was observed for 67 of the 87 diseases affected by higher LST, and for two of the three diseases affected by lower MVPA. |
| 13 | **Sensitivity analyses and additional analyses** | | | | |
| | | a) | Report any sensitivity analyses to assess the robustness of the main results to violations of the assumptions | 17-18 | Results of the sensitivity analyses are presented in Supplementary Table S6.<br>The effects identified in the discovery stage were consistent across sensitivity analyses (Supplementary Figure S2). The weighted median method supported the effects of higher LST, and genetic liability to lower MVPA, on the diseases identified in the discovery analyses. Most diseases (82/87 for higher LST; 6/6 for lower MVPA) showed no evidence of horizontal pleiotropy and the majority of MR-Egger estimates (64/87 for higher LST; 5/6 for lower MVPA) were consistent in direction with those obtained in the discovery analyses. |

| | | | | | |
|---|---|---|---|---|---|
| | | b) | Report results from other sensitivity analyses or additional analyses | 18 | Of the 87 diseases with potential effects of LST, UK Biobank GWAS data were available for 65; 51 of these replicated with an effect in the same direction and p ≤ 0.05, 11 showed directional consistency only and three showed no directional consistency. For lower MVPA, GWAS data were available for four of the six diseases; one replicated, and the remaining three showed directional consistency. Detailed results are provided in Supplementary Table S5. |
| | | c) | Report any assessment of direction of causal relationship (e.g., bidirectional MR) | 18 | Both higher LST, and genetic liability to lower MVPA, showed consistent effects on diseases after applying MR Steiger filtering. |
| | | d) | When relevant, report and compare with estimates from non-MR analyses | n/a | |
| | | e) | Consider additional plots to visualize results (e.g., leave-one-out analyses) | 18 | Supplementary Figure S2 |
| | **DISCUSSION** | | | | |
| 14 | **Key results** | | Summarize key results with reference to study objectives | 18-19 | In this large hypothesis-free MR-PheWAS, we found evidence of potential causal effects of sedentary behaviour (LST), with higher LST increasing the risk of 87 (5.1%) of the 1,719 diseases explored, most of which were in musculoskeletal and connective tissue, respiratory and genitourinary systems. Although we had lower power to detect effects of lower levels of physical activity (MVPA), the results were largely what we would expect given the correlation of this exposure with LST. We observed potential causal effects of increased risk of diseases in musculoskeletal and connective tissue, genitourinary and respiratory systems that were consistent with LST findings. In addition, of the 87 diseases that were significant for LST, 76 had estimated effects in the same direction for genetic liability to lower MVPA. Sensitivity analyses found all of these potential effects supported by weighted median method and most by MR-Egger. We were able to find GWAS for most of these potential effects for replication in UK Biobank, and 51 of the 65 effects were replicated, showing directional consistency and p ≤ 0.05, with 11 of the remainder being directionally consistent but having imprecise estimates. |

| | | | | |
|---|---|---|---|---|
| | | | | pain, sclerosis, etc.) and respiratory system (e.g., COPD, asthma, etc.) are consistent with findings from previous MR studies, whereas few such past studies have explored associations with genitourinary diseases (see Supplementary Table S1). Our finding of potential effects of LST and MVPA on increased risk of genitourinary diseases that have previously rarely been explored highlights the importance of hypothesis-free approaches that go beyond outcomes that are repeatedly studied.<br>Sedentary behaviour affected fewer cardiovascular diseases than reported in previous studies [43, 44], with the observed effects appearing to be primarily driven by hypertension, indicating that hypertension may be the cardiovascular disease most sensitive to sedentary behaviour. In relation to cardiovascular disease, RCTs of physical activity interventions have provided evidence that exercise-based cardiac rehabilitation reduces the risk of cardiovascular events [45, 46]. |
| | b) | Mechanism: Discuss underlying biological mechanisms that could drive a potential causal relationship between the investigated exposure and the outcome, and whether the gene-environment equivalence assumption is reasonable. Use causal language carefully, clarifying that IV estimates may provide causal effects only under certain assumptions | 19-20 | The diseases we identified as being potentially caused by high levels of sedentary behaviour and low physical activity, that were largely female reproductive conditions – such as cervix uteri, vagina or vulva conditions – suggest a possible role for physical activity in female pelvis strength, whereas the potential effect on menorrhagia may reflect the role of higher body mass index in polycystic ovary syndrome and reproductive hormones [39]. Conversely, it is possible that these findings reflect reverse causality (i.e. women with these conditions may be less able to exercise and thus more sedentary). However, Steiger filtering analyses suggested that these and other potential effects were unlikely due to reverse causality. Further research of how physical activity might affect women's genitourinary / reproductive system is warranted. |
| | c) | Clinical relevance: Discuss whether the results have clinical or public policy relevance, and to what extent they inform effect sizes of possible interventions | 21 | The adverse effects of higher sedentary behaviour and lower physical activity across multiple systems observed in this study indicate that interventions aimed at reducing sedentary behaviour and increasing moderate-to-vigorous physical activity could be beneficial for preventing multimorbidity. |

| | | | | |
|---|---|---|---|---|
| 17 | **Generalizability** | Discuss the generalizability of the study results (a) to other populations, (b) across other exposure periods/timings, and (c) across other levels of exposure | 22 | This study focused only on participants of European ancestry, which limits the generalizability of the findings to other populations with different genetic backgrounds. Studies based on larger sample sizes and non-European populations or trans-ancestry analyses are needed [51]. |
| | **OTHER INFORMATION** | | | |
| 18 | **Funding** | Describe sources of funding and the role of funders in the present study and, if applicable, sources of funding for the databases and original study or studies on which the present study is based | 25 | Funding<br>This study is funded by the European Union's Horizon Europe Research and Innovation Programme under grant agreement nº 101137146. UK participants in Horizon Europe Project STAGE are supported by UKRI grant numbers 10112787 (Beta Technology) and 10099041 (University of Bristol). RMAP's, GLC's and DAL's contribution was supported by the UK Medical Research Council and the University of Bristol (MC_UU_00032/2 and MC_UU_00032/5). DAL's contribution is also supported by the British Heart Foundation (CH/F/20/90003). |
| 19 | **Data and data sharing** | Provide the data used to perform all analyses or report where and how the data can be accessed, and reference these sources in the article. Provide the statistical code needed to reproduce the results in the article, or report whether the code is publicly accessible and if so, where | 24-25 | Availability of data and materials<br>FinnGen genetic summary statistics are available online (https://finngen.gitbook.io/documentation/r11/data-download). UK Biobank genetic summary data can be downloaded from the Neale lab (https://www.nealelab.is/uk-biobank), GeneATLAS (http://geneatlas.roslin.ed.ac.uk/downloads/) and OpenGWAS platform (https://gwas.mrcieu.ac.uk/datasets). Analysis plan and code for this study are available on Github (https://github.com/JiayaoXu2023/MR-PheWAS-SB-PA). |
| 20 | **Conflicts of Interest** | All authors should declare all potential conflicts of interest | 25 | Competing Interests<br>None |

This checklist is copyrighted by the Equator Network under the Creative Commons Attribution 3.0 Unported (CC BY 3.0) license.

1. Skrivankova VW, Richmond RC, Woolf BAR, Yarmolinsky J, Davies NM, Swanson SA, et al. Strengthening the Reporting of Observational Studies in Epidemiology using Mendelian Randomization (STROBE-MR) Statement. JAMA. 2021;under review.

2. Skrivankova VW, Richmond RC, Woolf BAR, Davies NM, Swanson SA, VanderWeele TJ, et al. Strengthening the Reporting of Observational Studies in Epidemiology using Mendelian Randomisation (STROBE-MR): Explanation and Elaboration. BMJ. 2021;375:n2233.
